# A Decision Analytics Model to Optimize Investment in Interventions Targeting the HIV PrEP Cascade of Care

**DOI:** 10.1101/2020.12.10.20247270

**Authors:** Samuel M. Jenness, Gregory Knowlton, Dawn K. Smith, Julia L. Marcus, Emeli J. Anderson, Aaron J. Siegler, Jeb Jones, Patrick S. Sullivan, Eva Enns

## Abstract

**Objectives:** Gaps between recommended and actual levels of HIV preexposure prophylaxis (PrEP) use remain among men who have sex with men (MSM). Interventions can address these gaps, but it is unknown how public health initiatives should invest prevention funds into these interventions to maximize their population impact.

**Design:** We used a stochastic network-based HIV transmission model for MSM in the Atlanta area paired with an economic budget optimization model.

**Methods:** The model simulated MSM participating in up to three real-world PrEP cascade interventions designed to improve initiation, adherence, or persistence. The primary outcome was infections averted over 10 years. The budget optimization model identified the investment combination under different budgets that maximized this outcome given intervention costs from a payer perspective.

**Results:** From the base 15% PrEP coverage level, the three interventions could increase coverage to 27%, resulting in 12.3% of infections averted over 10 years. Uptake of each intervention was interdependent: maximal use of the adherence and persistence interventions depended on new PrEP users generated by the initiation intervention. As the budget increased, optimal investment involved a mixture of the initiation and persistence interventions, but not the adherence intervention. If adherence intervention costs were halved, the optimal investment was roughly equal across interventions.

**Conclusions:** Investments into the PrEP cascade through initiatives should account for the interactions of the interventions as they are collectively deployed. Given current intervention efficacy estimates, the total population impact of each intervention may be improved with greater total budgets or reduced intervention costs.

## INTRODUCTION

Preexposure prophylaxis (PrEP) is highly effective at HIV prevention, but large gaps exist between actual and recommended levels of use [1]. Men who have sex with men (MSM) are a priority population for PrEP in the United States [2], but only 5–20% of MSM with indications currently use it [3]. The new *Ending the Epidemic* (EHE) plan seeks to reduce HIV incidence by 90% by 2030 [4]. Key to the EHE plan is the “Prevent” pillar, which includes efforts to close the gaps between recommended and actual levels of PrEP use among MSM in high-burden areas like Atlanta.

Opportunities to improve PrEP care have been characterized using a prevention cascade framework [5]. Steps in the PrEP cascade can be classified into three categories: initiation (including awareness, access, prescription), adherence, and persistence (i.e., retention in PrEP care). Several studies have been designed to evaluate behavioral and clinical interventions that target these steps in the PrEP cascade for MSM [6–8]. Achieving the EHE goals with PrEP will likely require a multi-pronged approach that improves all steps in the PrEP cascade. Understanding how combinations of initiation, adherence, and persistence interventions scale and interact is needed to direct EHE and local investment priorities.

Transmission modeling of the PrEP cascade suggests that improving initiation and persistence are needed to further advance HIV prevention [9,10]. However, modeling studies to date have represented cascade improvements non-mechanistically: models projected what happens when rates of PrEP initiation increase and rates of discontinuation decrease, but not the mechanisms by which those changes could occur. These modifications to the PrEP cascade may require investments in activities involving health system change (e.g., telemedicine) and targeted interventions using both traditional (e.g., counseling) or novel (e.g., mobile apps) approaches. Because these activities require financial resources, health authorities must consider how to invest EHE and other funding to balance the epidemiological benefits of these interventions against their costs.

In this study, we used decision analytic modeling pairing a stochastic network-based model of HIV transmission dynamics among MSM with a budget optimization model. These models simulated HIV transmission under varying levels of engagement in three categories of interventions (initiation, adherence, persistence) reflecting three real-world tools with unique impact on the PrEP cascade. Our research objective was to determine the optimal investment of a dedicated budget across these three categories of interventions to maximize the expected number of infections averted over 10 years.

## METHODS

### Study Design

Our network-based model of HIV transmission dynamics was built with the EpiModel platform [11]. Building on our previous models [12], this study implemented the mechanisms for increasing PrEP coverage through engagement in interventions. Full methodological details are provided in an Appendix [**LINK; Supplemental Tables 1–13; Supplemental Figures 1–8**], and full model code is available at http://github.com/EpiModel/PrEP-Optimize. The model represented main, casual, and one-time sexual partnerships for Black, Hispanic, and White/Other MSM, aged 15 to 65, in the Atlanta area. The starting network size in the model was 10000 MSM, which stochastically fluctuated over time with arrival (sexual debut) and departure (mortality or sexual cessation) (**Appendix S5**).

### HIV Transmission and Progression

The epidemic model consisted of five linked components: 1) statistical network models to generate dynamic sexual partnerships; 2) statistical models to predict behavior within partnerships; 3) simulation of HIV transmission in partnerships; 4) simulation of HIV progression and other natural history features; and 5) simulation of prevention and treatment services.

To fit the network models, we used data from ARTnet, an egocentric network study conducted in 2017–2019 of US MSM [13] (**Appendix S2**). Parameters were weighted by census-based race/ethnicity and age distributions to account for sampling biases. Predictors of partnership formation included partnership type, degree distributions by partnership types, heterogeneity in degree and assortative mixing by race/ethnicity, age, and mixing by sexual position (**Appendix S3**). Partnership durations were modeled with dissolution rates stratified by partnership type and age. Models were then fit to ARTnet to predict coital frequency and condom use probability as a function of race/ethnicity, age, diagnosed HIV status, and partnership type and duration (**Appendix S4**).

In each weekly time step, MSM could be screened for HIV. MSM who screened positive entered the HIV care cascade (**Appendix S7**). MSM linked to care and initiating antiretroviral therapy (ART) had reduced vial load (VL) and increased longevity. MSM on ART could cycle off and back on ART based on rates calibrated to local surveillance data [14]. Lower VL with sustained ART use was associated with a reduced probability of HIV transmission per act. Other factors modifying the HIV transmission probability included PrEP use, condom use, sexual position, and circumcision (**Appendix S8**).

### Baseline PrEP Cascade

HIV screening was also the entry point to PrEP [5]. MSM screening negative entered the PrEP cascade if they met indications based on CDC guidelines [2]. Baseline PrEP care was parameterized to reflect PrEP engagement observed in the Atlanta area (**Table 1**). Indicated MSM initiated PrEP at a probability generating a coverage level (percent of indicated MSM on PrEP) of 15% [15]. Heterogeneous PrEP adherence was represented as two levels (high versus medium/low), which corresponded to different relative reductions in HIV acquisition risk [16,17]. Spontaneous PrEP discontinuation was based on data for the proportion of MSM who were retained in PrEP care at 6 months [18]. MSM also stopped PrEP if they no longer exhibited PrEP indications [2].

**Table 1.**
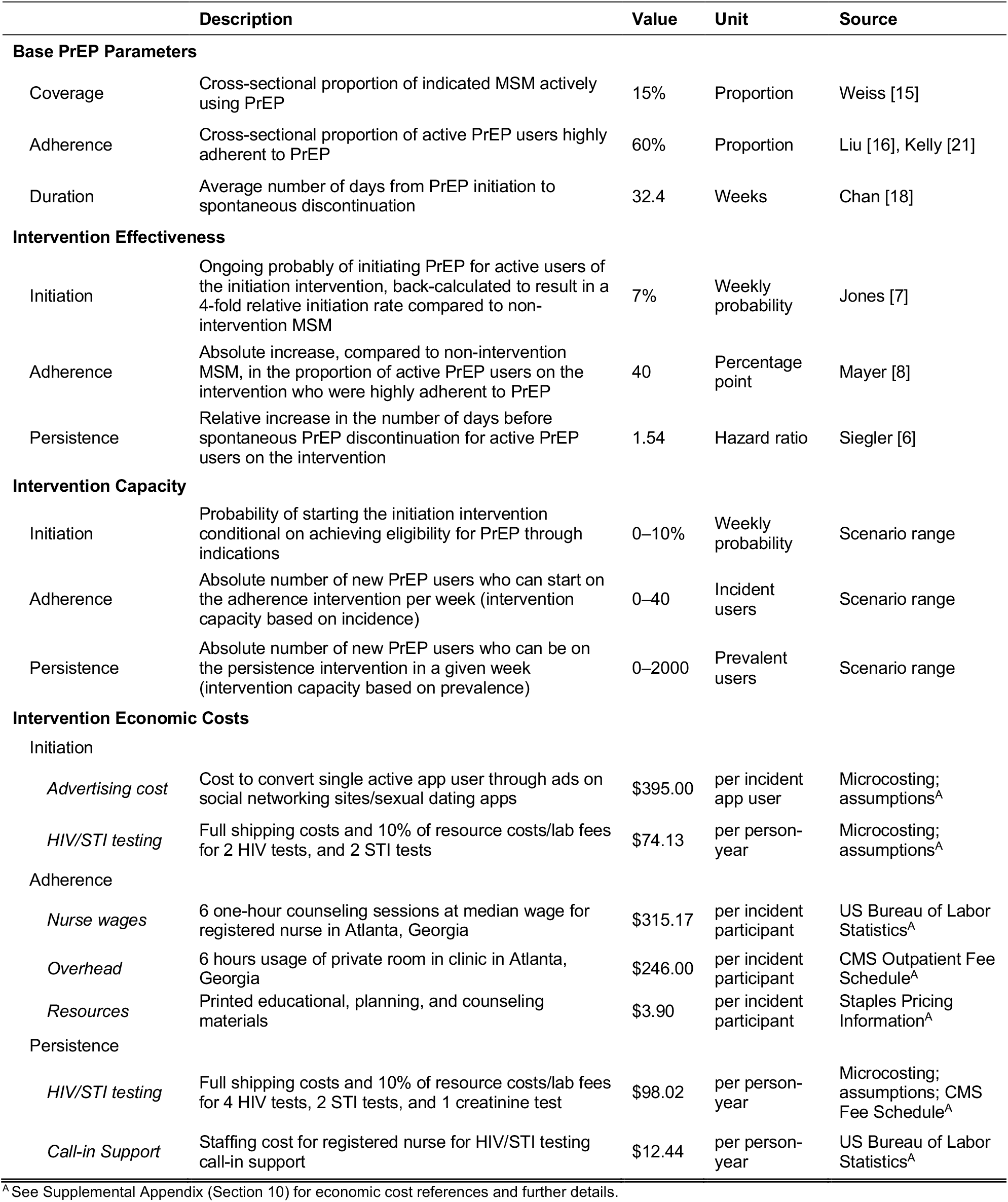
Epidemiological and Economic Model Parameters

|  | Description | Value | Unit | Source |
| --- | --- | --- | --- | --- |
| **Base PrEP Parameters** |  |  |  |  |
| Coverage | Cross-sectional proportion of indicated MSM actively using PrEP | 15% | Proportion | Weiss [15] |
| Adherence | Cross-sectional proportion of active PrEP users highly adherent to PrEP | 60% | Proportion | Liu [16], Kelly [21] |
| Duration | Average number of days from PrEP initiation to spontaneous discontinuation | 32.4 | Weeks | Chan [18] |
| **Intervention Effectiveness** |  |  |  |  |
| Initiation | Ongoing probability of initiating PrEP for active users of the initiation intervention, back-calculated to result in a 4-fold relative initiation rate compared to non-intervention MSM | 7% | Weekly probability | Jones [7] |
| Adherence | Absolute increase, compared to non-intervention MSM, in the proportion of active PrEP users on the intervention who were highly adherent to PrEP | 40 | Percentage point | Mayer [8] |
| Persistence | Relative increase in the number of days before spontaneous PrEP discontinuation for active PrEP users on the intervention | 1.54 | Hazard ratio | Siegler [6] |
| **Intervention Capacity** |  |  |  |  |
| Initiation | Probability of starting the initiation intervention conditional on achieving eligibility for PrEP through indications | 0–10% | Weekly probability | Scenario range |
| Adherence | Absolute number of new PrEP users who can start on the adherence intervention per week (intervention capacity based on incidence) | 0–40 | Incident users | Scenario range |
| Persistence | Absolute number of new PrEP users who can be on the persistence intervention in a given week (intervention capacity based on prevalence) | 0–2000 | Prevalent users | Scenario range |
| **Intervention Economic Costs** |  |  |  |  |
| Initiation |  |  |  |  |
| *Advertising cost* | Cost to convert single active app user through ads on social networking sites/sexual dating apps | \$395.00 | per incident app user | Microcosting; assumptions<sup>A</sup> |
| *HIV/STI testing* | Full shipping costs and 10% of resource costs/lab fees for 2 HIV tests, and 2 STI tests | \$74.13 | per person-year | Microcosting; assumptions<sup>A</sup> |
| Adherence |  |  |  |  |
| *Nurse wages* | 6 one-hour counseling sessions at median wage for registered nurse in Atlanta, Georgia | \$315.17 | per incident participant | US Bureau of Labor Statistics<sup>A</sup> |
| *Overhead* | 6 hours usage of private room in clinic in Atlanta, Georgia | \$246.00 | per incident participant | CMS Outpatient Fee Schedule<sup>A</sup> |
| *Resources* | Printed educational, planning, and counseling materials | \$3.90 | per incident participant | Staples Pricing Information<sup>A</sup> |
| Persistence |  |  |  |  |
| *HIV/STI testing* | Full shipping costs and 10% of resource costs/lab fees for 4 HIV tests, 2 STI tests, and 1 creatinine test | \$98.02 | per person-year | Microcosting; assumptions; CMS Fee Schedule<sup>A</sup> |
| *Call-in Support* | Staffing cost for registered nurse for HIV/STI testing call-in support | \$12.44 | per person-year | US Bureau of Labor Statistics<sup>A</sup> |

<sup>A</sup> See Supplemental Appendix (Section 10) for economic cost references and further details.

*PrEP Cascade Interventions*. With the calibrated model, we then simulated three categories of PrEP interventions at different levels of capacity. MSM could join one or more interventions, subject to capacity constraints. Engagement in each of the three interventions was which was associated with improvements to in PrEP initiation, adherence, or persistence. MSM not on an intervention continued *status quo* PrEP care in that domain.

The initiation intervention was modeled after **Healthmindr**, currently in clinical trials [7]. This intervention features a phone app delivering information and resources about PrEP, including individualized risk assessments and PrEP provider locations. The goal of the Healthmindr intervention is to increase the rate of initiation for PrEP-indicated MSM. In our model, MSM who were indicated for but not using PrEP were eligible to start Healthmindr, with a range of app initiation probabilities that translated to different levels of Healthmindr capacity (**Table 1**). MSM on Healthmindr then had an additional PrEP initiation probability (elevating the baseline probability) that corresponded to the intervention efficacy, which was based on power analyses in the study protocol [7]. MSM dropped off the intervention when indications lapsed, but they could reenter Healthmindr upon new indications.

The adherence intervention was modeled after **Life-Steps for PrEP**, completed in 2017 [8]. Life-Steps used cognitive behavioral counseling delivered by nurses to improve medication adherence for MSM currently using PrEP. The trial found that PrEP drug levels were significantly higher in the intervention arm. In our model, MSM were eligible for Life-Steps at PrEP initiation. The intervention had a fixed number of new users each week, which we varied up to the maximum service capacity. This intervention had the effect of shifting the probability of assignment into the high adherence category from 60% (baseline) to 100% for those enrolled. This was an intervention with effects lasting the duration of a PrEP episode. A PrEP episode is one instance of starting and (potentially) stopping PrEP, and one person may have multiple episodes; the intervention effects from one episode did not impact adherence in future episodes.

The persistence intervention was modeled after **ePrEP** and **PrEP@Home** interventions, currently in trials [6,19]. Both feature home-based PrEP care accomplished through a mobile phone app, specimen self-collection, and video-based clinical consultations. Primary intervention outcomes measure persistence on PrEP after initiating the intervention. In our model, MSM were eligible for the persistence intervention at the point of PrEP initiation. The intervention had a fixed capacity of prevalent (active) users, which we varied up to the maximum clinical service capacity expected if the intervention were fully scaled up. Intervention participants had a lower rate of spontaneous PrEP discontinuation compared to non-intervention PrEP users. This lower rate was informed by power analyses in the protocol [6]. PrEP users initiating ePrEP/PrEP@Home were assumed to stay on the intervention for the full duration of their PrEP episode. Thus, MSM stopped the intervention only if they discontinued PrEP, either spontaneously or through lapsed indications.

### Epidemic Model Calibration and Simulation

We calibrated the epidemic model with a Bayesian approach that defined prior distributions for uncertain parameters and fit the model to empirical surveillance-based estimates for the target population (**Appendix S9**). After calibration, we randomly drew from a multivariate distribution of capacity parameters for the three interventions (**Table 1**) using Latin hypercube sampling with uniform probability distributions. This generated 1000 capacity parameter sets. For each set, we then simulated the epidemic model 250 times for 10 years in weekly time steps. The primary outcome was the median cumulative 10-year incidence across the 250 simulations. We compared the median cumulative incidence in each of the 1000 sampled parameter sets against that in the base scenario (no PrEP cascade interventions).

### Budget Optimization Model

A nonlinear programming (NLP) model was used to solve for the optimal allocation of the budget for these three interventions, maximizing infections averted. The NLP was defined by a nonlinear objective function, a linear constraint related to the total budget, and bounds for the minimum and maximum allowable intervention capacities. The optimization can be represented mathematically as:

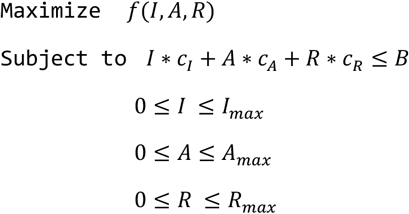

where *f*() is the objective function, *c*_*I,A,R*_ are the unit costs associated with the interventions for the entire 10-year time horizon, and *B* is the 10-year budget allocated to improving the PrEP cascade. The objective function is the expected infections averted over the 10-year time horizon as a function of the clinical capacities of the initiation (*I*), adherence (*A*), and persistence (*R*) interventions. In terms of capacity measures, *I* is the percentage of newly indicated MSM reached by the initiation intervention, *A* is the number of new PrEP users allowed to start the adherence intervention, and *R* is the maximum number of PrEP users engaged in the persistence intervention at any time. We estimated the objective function through a generalized additive model fit to the simulated infections averted over the considered ranges of intervention capacities (**Appendix S10**).

For all three interventions, we assumed that the intervention costs increased linearly with capacity. The budget optimization was conducted from a payer-perspective, so only costs that would be paid by a hypothetical federal EHE or local health jurisdiction budget allocated to PrEP improvement were included. For each intervention, we estimated the resources needed to administer the intervention per person or person-time and then estimated unit costs of each resource type, using Atlanta-specific estimates when available (**Table 1**). For the initiation intervention, costs included the advertising needed to attract new PrEP users. These intervention costs also included 10% of the HIV/STI screening resources and laboratory costs incurred by users (the estimated proportion of these costs that would be covered directly by the budget based on the local uninsured rate) [20]. Adherence intervention costs included personnel and physical space to administer the intervention.

## RESULTS

**Figure 1** shows the impact of scaling up the capacity of the initiation and persistence interventions on PrEP coverage (**Panel A**) and HIV incidence (**Panel B**). In general terms, the maximum initiation intervention capacity was roughly equal to the current size of the PrEP-eligible population, whereas the maximum adherence and persistence capacity levels were roughly equal to the current size of the prevalent PrEP user base. Scaling up the initiation and persistence interventions to their maximum considered capacities resulted in PrEP use among 27% of indicated MSM, an increase of 12 percentage points from the baseline PrEP coverage of 15%. Higher capacity on both the initiation and persistence interventions increased PrEP coverage, but PrEP coverage improvements from scaling up capacity in the persistence intervention were constrained by the capacity of the initiation intervention. PrEP coverage as impacted by the persistence intervention capacity plateaued when there was capacity to provide nearly all prevalent PrEP users with the persistence intervention. Additional persistence intervention capacity provided no benefit beyond this point (if all prevalent PrEP users are reached by the persistence intervention, then any additional investment into the persistence intervention does not generate any additional benefit). Before this plateau was reached, however, the marginal effect of scaling up the persistence intervention was similar across different levels of initiation intervention scale-up. This persistence intervention capacity was not a fixed number across the scenarios but depended on the capacity of the initiation intervention as it increased incident PrEP use. Because of the model stochasticity, the persistence intervention use fluctuated relative to capacity (there were some weeks when that capacity was used and some when it was not). **Panel B** shows how the percent infections averted changes as a function of these different combinations of initiation and persistence capacities. When both interventions were scaled up to their maximum capacities, there was a 9.3% reduction in cumulative incidence.

**Figure 1.**
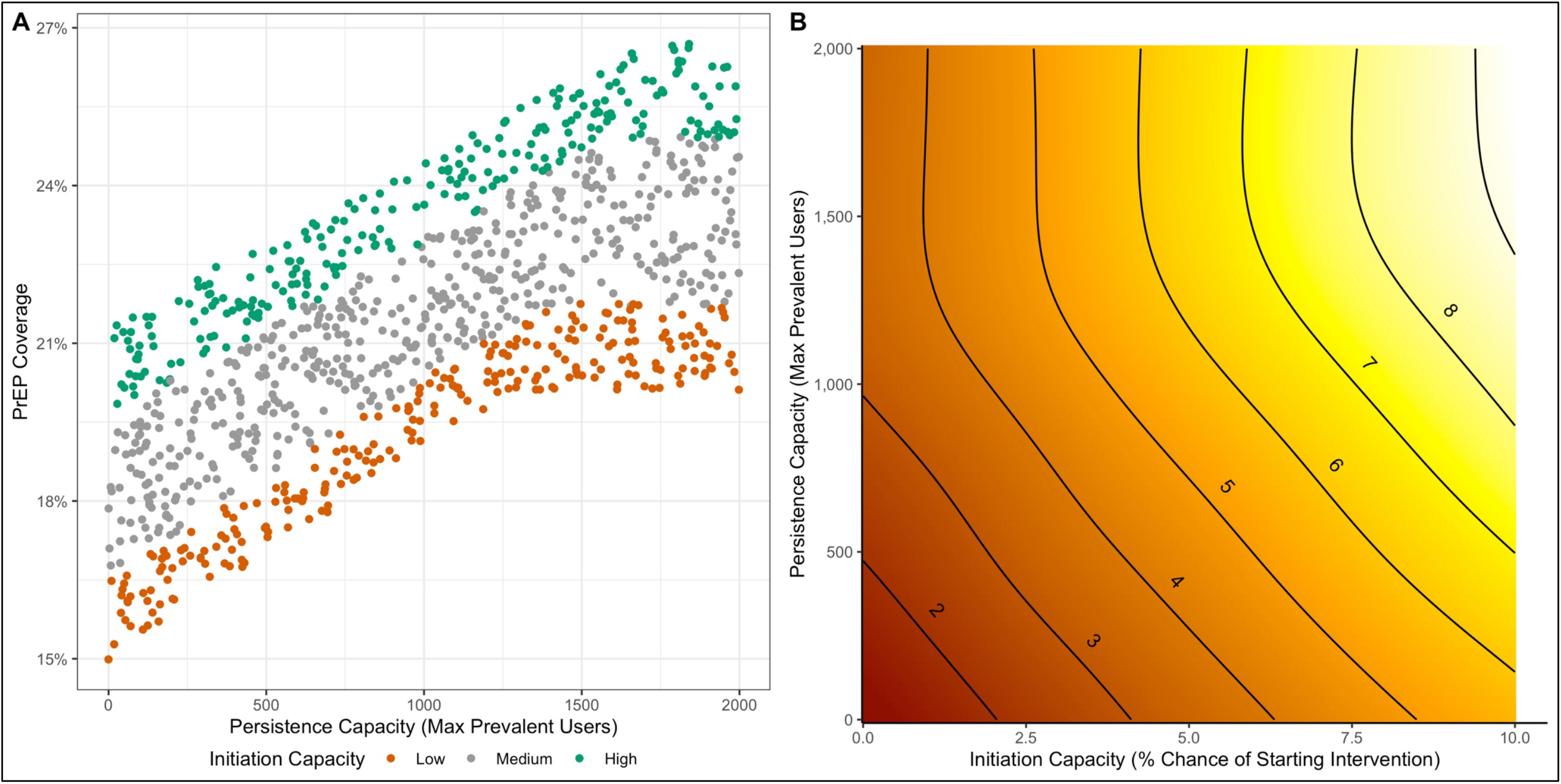
Panel A shows the relationship between capacity on initiation (probability of starting initiation intervention) and capacity on persistence interventions on PrEP coverage. Each dot represents one parameter scenario set (medians across 250 individual simulations). Panel B shows the relationship of the initiation and persistence capacity on the percent of cumulative infections averted over 10 years (integer values next to the bands), with lighter colors indicating more infections inverted due to the two interventions in tandem.

**Figure 2** demonstrates the joint interaction of the three interventions. Maximizing capacity of all three interventions averted 12.3% of HIV infections compared to the base scenario. The marginal increase in infections averted with increasing persistence intervention capacity was independent of the adherence intervention capacity, except when PrEP initiation was low. When the initiation intervention capacity was low, a unit increase in persistence capacity provided roughly the same benefit for both medium and high adherence intervention capacities. However, for medium-to-high levels of initiation intervention capacity, increasing adherence intervention capacity had the impact of shifting the overall persistence intervention response curve higher. Overall, the more person-time on PrEP generated through initiation and persistence interventions, the more valuable it was to make that person-time highly adherent.

**Figure 2.**
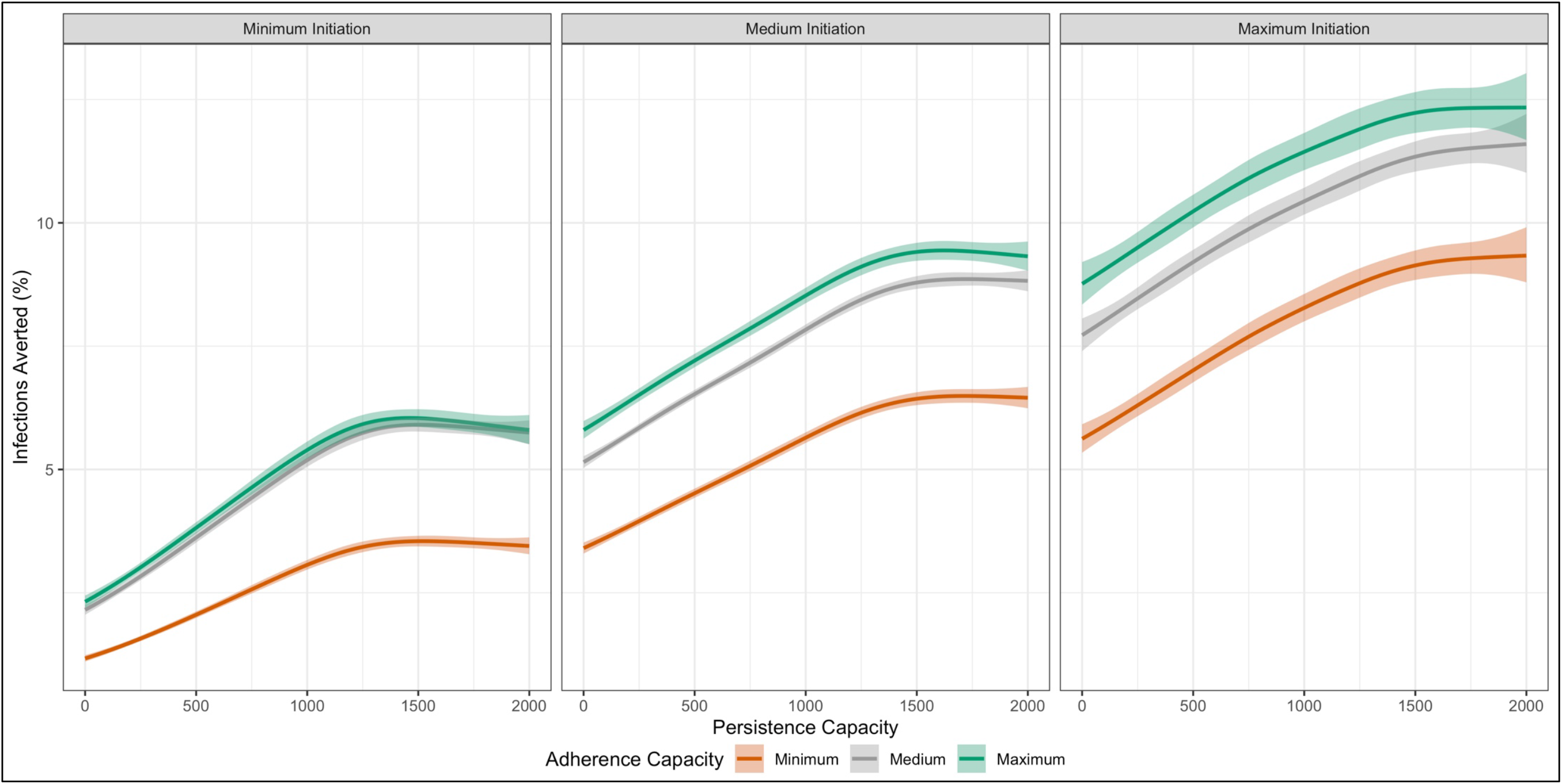
Three panels show the impact of variations in three interventions together on 10-year cumulative infections averted. Subpanels show minimum, medium (median), and maximum capacity for the initiation intervention, where the individual lines in each panel show minimum, medium, and maximum capacities of the adherence intervention (same definitions as intervention capacity). Thick lines are the point estimates from the GAMs fit to the mean of the simulations; bands represent 95% confidence intervals around those point estimates.

**Figure 3** (**Panels A and B**) and **Table 2** present the results of the model of investment strategies across 10-year budget levels that maximize infections averted. The optimal strategy involved a mixture of initiation and persistence interventions investment across all budget levels. At no budget level was it considered optimal to invest into the adherence intervention. At lower budgets (less than $1.45 million), investment entirely in the persistence intervention was optimal. As the budget increased, a mixture of investments in the initiation and persistence interventions achieved the greatest benefits. As **Panel B** shows, higher levels of investment in the initiation intervention did not displace the budget allocated to the persistence intervention. Instead, the optimal budget allocation was to expand the persistence intervention capacity to fully meet the demand from MSM on PrEP who could utilize it; once that demand was satisfied, any remaining funds were allocated to increase initiation. At the highest 10-year budget level ($6 million), the optimal allocation was to invest 26% of the budget into the persistence intervention, while investing the remaining budget into initiation. At this budget level and investment strategy, PrEP coverage would increase to 24% (from 15% in the base model), averting 76.5 (6.4%) HIV infections over 10 years. The intervention cost per HIV infection averted increased with the total budget, ranging from $25,048 at the lowest budget to $69,281 for the highest total budget considered, indicating diminishing returns as the budget was increased.

**Table 2.** Optimal Allocation to Three PrEP Cascade Interventions (Initiation, Adherence, Persistence) across Different 10-Year Budgets to Minimize Cumulative HIV Incidence over 10 Years (Primary Budget Scenarios), Relative to No Intervention Investment (Base Scenario)

| Outcomes | 10-Year Budget Cap |  |  |  |  |  |
| --- | --- | --- | --- | --- | --- | --- |
| | \$0 (Base) | \$700,000 | \$1,500,000 | \$3,000,000 | \$4,500,000 | \$6,000,000 |
| **Optimal Intervention Value (Means)** |  |  |  |  |  |  |
| Initiation Probability (%) | – | 0 | 0.1 | 2.3 | 4.3 | 6.3 |
| Adherence Capacity (N) | – | 0 | 0 | 0 | 0 | 0 |
| Persistence Capacity (N) | – | 634 | 1300 | 1295 | 1382 | 1460 |
| **Total Spending by Intervention (Means)** |  |  |  |  |  |  |
| Initiation (\$, in thousands) | 0 | 0 | 64 | 1570 | 2973 | 4387 |
| Adherence (\$, in thousands) | 0 | 0 | 0 | 0 | 0 | 0 |
| Persistence (\$, in thousands) | 0 | 700 | 1436 | 1430 | 1527 | 1613 |
| **Proportional Spending by Intervention (Means)** |  |  |  |  |  |  |
| Initiation (%) | 0.0 | 0.0 | 4.3 | 52.3 | 66.1 | 73.1 |
| Adherence (%) | 0.0 | 0.0 | 0.0 | 0.0 | 0.0 | 0.0 |
| Persistence (%) | 0.0 | 100.0 | 95.7 | 47.7 | 33.9 | 26.9 |
| **Process Outcomes (Medians, 95% SI<sup>A</sup>)** |  |  |  |  |  |  |
| PrEP Coverage (%) | 15.0 (14.2, 15.9) | 17.5 (16.6, 18.4) | 20.1 (19.2, 21.0) | 21.3 (20.4, 22.2) | 22.7 (21.8, 23.6) | 24.0 (23.2, 25.0) |
| PrEP Highly Adherent (%) | 59.9 (57.2, 63.1) | 60.0 (57.5, 63.0) | 60.0 (57.5, 62.4) | 60.3 (57.6, 62.8) | 60.4 (57.8, 62.9) | 60.3 (57.9, 62.6) |
| PrEP Discontinuation Rate<sup>B</sup> | 2.2 (1.3, 3.3) | 1.7 (1.0, 2.5) | 1.4 (0.8, 2.0) | 1.4 (0.9, 2.0) | 1.5 (0.9, 2.1) | 1.5 (0.9, 2.1) |
| **Epidemiological Outcomes (Medians, 95% SI)** |  |  |  |  |  |  |
| HIV Incidence Rate<sup>C</sup> | 1.30 (1.02, 1.64) | 1.28 (1.02, 1.58) | 1.24 (0.94, 1.56) | 1.23 (0.94, 1.55) | 1.21 (0.97, 1.52) | 1.19 (0.92, 1.45) |
| Cumulative HIV Incidence<sup>D</sup> | 1194 (1105, 1296) | 1174 (1090, 1268) | 1160 (1063, 1254) | 1146 (1052, 1230) | 1127 (1031, 1214) | 1118 (1034, 1210) |
| Number Infections Averted | – | 20.0 (14.0, 26.0) | 35.0 (29.0, 41.0) | 48.5 (42.5, 54.0) | 65.0 (59.0, 71.0) | 76.5 (70.0, 82.0) |
| Percent Infections Averted | – | 1.7 (1.2, 2.1) | 2.9 (2.5, 3.4) | 4.1 (3.6, 4.5) | 5.5 (4.9, 5.9) | 6.4 (5.9, 6.9) |
| **Economic Outcomes (Means, 95% CI)** |  |  |  |  |  |  |
| Cost Per Infection Averted (\$) | – | 25048<br>(24309, 25866) | 35613<br>(34546, 36627) | 53620<br>(52605, 54602) | 63453<br>(62198, 64593) | 69281<br>(68003, 70663) |

<sup>A</sup> SI: simulation interval (middle 95% of simulated values) <sup>B</sup> Rate per 100 active PrEP users per week. <sup>C</sup> Incidence rate of HIV per 100 person-years at risk, averaged across the final year of the simulation (year 9); Rate and 95% SI. <sup>D</sup> Cumulative incidence in simulated population across 10 years; Count and 95% SI.

**Figure 3.**
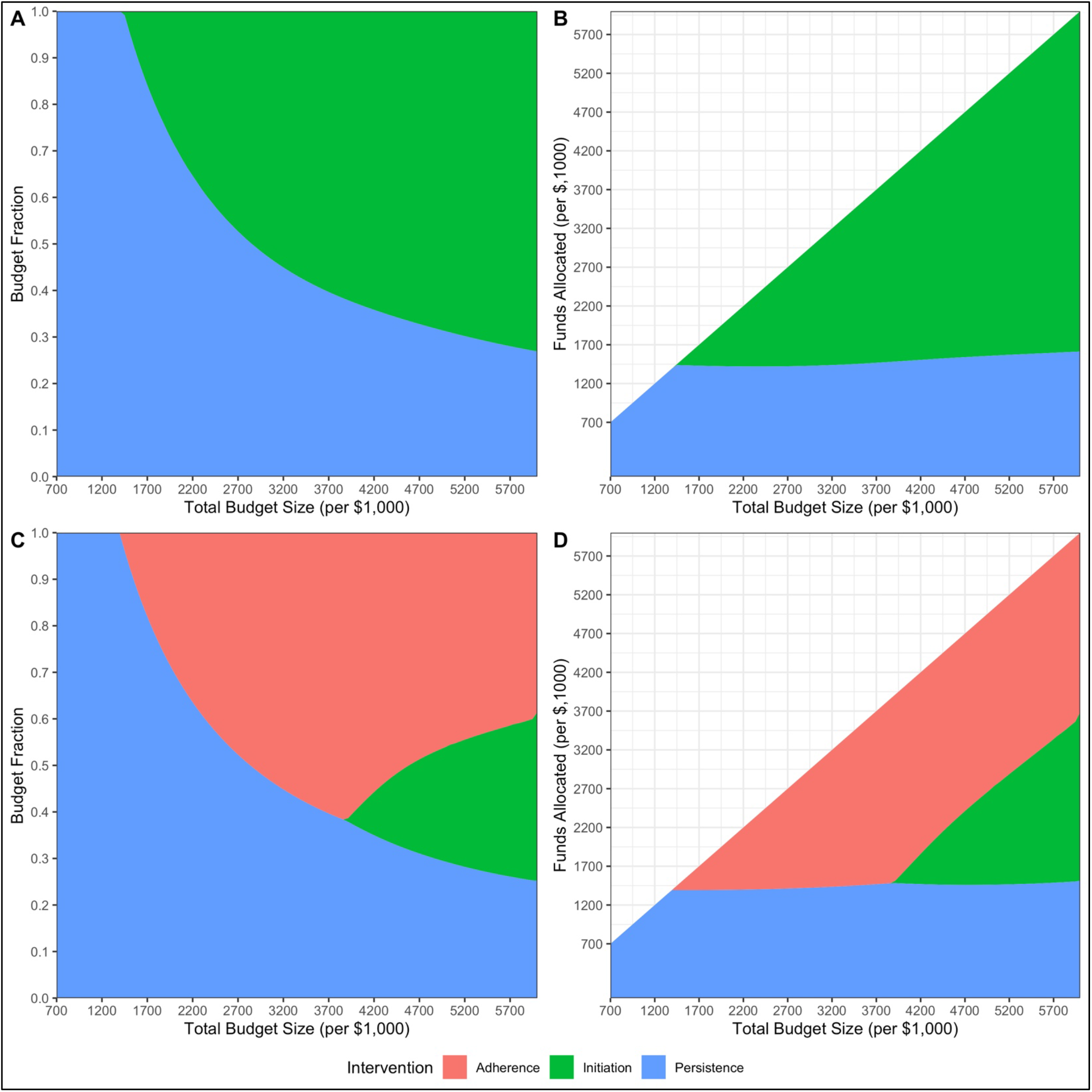
Results of the optimization analyses on budgetary allocation to each of the three interventions over different total 10-year budget sizes. Panels A and B show the fractional and absolute funds of the budget allocated to each intervention category in the primary (empirical) unit costs. Panels C and D show the same for the sensitivity optimization scenarios in which the unit costs of the adherence intervention are reduced by approximately half (55%).

Despite having an epidemiological benefit on HIV incidence (**Figure 2**), the adherence intervention was not selected for investment in the primary budget optimization model. The reason was its higher relative costs per infection averted compared to the initiation and persistence interventions. This was driven by the higher unit costs associated with nurse personnel time for behavioral counseling in that intervention. In a sensitivity analysis (**Figure 3, Panels C and D**, and **Supplemental Table 14**), we explored how reductions in the costs for this intervention (e.g., by having the counseling conducted by non-nurse staff) would impact the budget optimization. When the adherence intervention unit costs were reduced by approximately half (55%), the adherence intervention became a favorable investment. The persistence intervention still absorbed all of the funding at the lowest budget levels. However, unlike with the primary optimization model, when the persistence capacity was met, the adherence intervention was first selected for investment over the initiation intervention. The adherence intervention then exhibited a diminishing return at middle budget sizes (approximately $3.9 million for 10 years), at which point some investment in the initiation intervention was optimal. At the highest budget levels, the distribution of spending for the initiation, adherence, and persistence interventions was roughly equal (35%, 40%, and 25%, respectively).

## DISCUSSION

This study investigated how to optimally allocate funds across three categories of interventions targeting gaps in the PrEP cascade. This was the first model using a budget optimization approach for PrEP investment. We found that with increasing budgets, the persistence intervention should first be funded until its capacity fully satisfies user demand. At that point, the initiation intervention should be funded to increase overall PrEP coverage, and to further grow demand for the persistence intervention. Investments in the adherence intervention were not optimal based on current estimated unit costs; if those costs were reduced by half, the optimal allocation would fund three interventions at roughly equal levels. This study highlights that EHE and related investments into the PrEP cascade should account for the interacting impacts of different interventions as they are deployed.

Despite progress in delivery of PrEP to populations in need, persistent gaps in the PrEP cascade limit its optimal use [21]. Our model estimated the optimal distribution of investments in interventions to close these gaps, given the epidemiology of an EHE target jurisdiction [1]. EHE investments have the effect of changing the current baseline conditions in meaningful ways. Each level of increasing investment in these interventions effectively shifts the baseline to a better level. Further investment increases the overall infections averted but does so at a higher cost per infection averted.

Our study demonstrates the importance of understanding the mechanisms for how to close these PrEP cascade gaps. An unanswered implementation science question is why weaker than projected prevention benefits are seen as PrEP is scaled up [22]. One analysis found only modest declines in HIV diagnoses in states with higher PrEP prescription rates [23]. While this may be partially due to diagnoses being an imperfect proxy for incidence, PrEP prescriptions do not necessarily translate into optimal levels of PrEP use [24]. Retaining MSM in PrEP care continues to be a major challenge [25], and MSM are at much higher HIV risk immediately after discontinuing PrEP than while on PrEP [26]. Our model suggests that spending on initiation is optimal only once PrEP persistence gaps are closed. The benefits of generating new PrEP prescriptions (e.g., with the initiation intervention) will not translate into maximum prevention benefits unless those new users are retained on PrEP during their sexual risk period. Although the adherence intervention also had an epidemiological impact, reducing its unit costs would be required for selection into an optimal investment mix. This might be possible by having peer educators rather than nurses deliver the counseling sessions, but the effectiveness of that approach has not been evaluated.

Our representation of the PrEP cascade mechanisms drives the methodological innovation of this study. Many HIV economic models have used cost-effectiveness analysis (CEA) approaches to balance the financial costs against the epidemiological benefits of PrEP [27,28]. However, CEAs primarily focus on the efficiency of PrEP itself and assume it can be scaled up to high levels of coverage; CEAs of PrEP have not, to date, accounted for the resources needed to achieve those levels of scale-up and to support high levels of adherence and persistence. This is an illustration of a common bias in CEAs to ignore the cost of the supportive activities needed to ensure the success of biomedical interventions. Furthermore, CEAs can inform whether an HIV prevention strategy, such as PrEP, achieve health benefits at a reasonable level of health sector efficiency. However, CEAs do not inform whether a given intervention should be adopted by a given payer, especially when that payer is subject to substantially smaller budgets than the entirety of US health care spending. Our budget optimization approach, in contrast, takes a direct payer perspective and can therefore inform how the CDC or local health jurisdictions should spend funds on services to amplify PrEP benefits in order to maximize HIV prevention benefits. As the price of PrEP medication decreases through generic TDF/FTC [29] or subsidies from the *Ready, Set, PrEP* program, the costs of ancillary clinical services (e.g., routine STI screening in PrEP care [30]) and interventions to close the gaps in the PrEP cascade will constitute a larger portion of PrEP-related spending. Thus, financial considerations for PrEP programs will shift from financing medication access to funding initiatives that support optimal PrEP use. Nonetheless, if PrEP medication and other healthcare costs were included in our optimization model, the optimal selection of interventions would likely shift away from those that increased coverage (initiation and persistence interventions) and towards the adherence intervention (which maximizes efficiency of medication). We provide additional considerations in **Appendix S10**.

### Limitations

The primary limitation of our study is the assumptions about the individual-level efficacy of the three modeled interventions. For Healthmindr and ePrEP/PrEP@Home, efficacy has not been precisely determined because their clinical trials are incomplete. However, our efficacy estimates were drawn from power analyses in the trial protocols, which themselves were based on pilot data on the expected efficacy. We expect our results would apply to other interventions with demonstrated efficacy similar our assumed values. Second, we made our best effort to quantify the unit costs for these interventions that would likely be paid by EHE or local funds if the interventions were deployed. This required some cost assumptions (e.g., online advertising costs needed to generate a new PrEP user). Formal costing studies are also underway for the ongoing trials that could change these selected costs. Finally, our results may not be transportable to settings with different baseline HIV incidence or PrEP coverage. Our target population of Atlanta MSM was selected based on its reflection of many EHE target jurisdictions precisely where these PrEP cascade interventions are needed. However, we would expect PrEP coverage and HIV incidence to have predictable results: If PrEP coverage were higher, the persistence intervention would be efficient at even higher capacity levels and the switch to the initiation intervention would occur at higher relative budgets; if PrEP coverage were lower, the capacity ceiling for the persistence intervention would be hit sooner and the switch to the initiation intervention would happen at lower budgets; if PrEP coverage were lower and HIV incidence higher, the model would likely suggest expanding new PrEP users first (through additional investment in the initiation intervention) up to some critical level.

### Conclusions

Closing the gaps between actual and recommended PrEP use will depend on deploying interventions that address the steps of the PrEP cascade. Our study highlights the importance of understanding the mechanistic synergies of these interventions when optimizing these investments within HIV prevention initiatives like the EHE.

## Data Availability

The model code and parameters are all open and availability at the Github repository linked below.

https://github.com/EpiModel/PrEP-Optimize

